# Omnibus proteome-wide association study (PWAS-O) identified 43 risk genes for Alzheimer’s disease dementia

**DOI:** 10.1101/2022.12.25.22283936

**Authors:** Tingyang Hu, Randy L. Parrish, Qile Dai, Aron S. Buchman, Shinya Tasaki, David A. Bennett, Nicholas T. Seyfried, Michael P. Epstein, Jingjing Yang

## Abstract

Proteome-wide association study (PWAS) integrating proteomics data with GWAS data is a powerful tool to identify risk genes for complex diseases, which can inform disease mechanisms with genetic effects mediated through protein abundance. We propose a novel omnibus method to improve PWAS power by modeling unknown genetic architectures with multiple statistical models. We applied TIGAR, PrediXcan, and FUSION to train protein abundance imputation models for 8,430 proteins from dorsolateral prefrontal cortex with whole genome sequencing data (n=355). Next, the trained models were integrated with GWAS summary data of Alzheimer’s disease (AD) dementia (n=762,917) to conduct PWAS. Last, we employed the Aggregated Cauchy Association Test to obtain omnibus PWAS (PWAS-O) p-values from these three models. PWAS-O identified 43 risk genes of AD dementia including 5 novel risk genes that were interconnected through a protein-protein interaction network including *TOMM40*, *APOC1*, and *APOC2*. PWAS-O can be easily applied to study complex diseases.

## Introduction

Recent large-scale genome-wide association studies (GWAS) of Alzheimer’s disease (AD) dementia have identified 38 risk loci^1^, but little is known about their underlying mechanisms. Integrating transcriptomic data with GWAS data through transcriptome-wide association study (TWAS) has been shown an effective approach for identifying potential causal risk genes of complex human diseases with biological interpretations^2–4^. TWAS can inform potential underlying disease mechanisms as the identified genetic effects may be potentially mediated via gene expression^5, 6^. Since many biological processes can affect protein abundance, gene expression levels might not be the optimal proxy for elucidating the role of proteins in disease mechanisms. The analogous proteome-wide association study (PWAS) approach integrating proteomic data with GWAS summary data^7–9^ has the potential to identify novel genetic risk loci linked to disease via proteins that may be missed by GWAS and TWAS^2–4, 10^.

Since gene expression and protein abundance are both molecular quantitative traits that might mediate genetic effects, existing TWAS tools can be used for PWAS. TWAS tools can train protein abundance imputation models using genetic variants as predictors from a reference panel with both proteomic and genetic data available. Analogously, genetic effect sizes in the imputation models could be viewed as effect sizes of “protein quantitative trait locus (pQTL)” in a broad sense (as these “pQTL” are not necessarily statistically significant), which would be taken as variant weights for gene-based association study (i.e., PWAS) with GWAS data.

Existing TWAS tools use various statistical regression models to train imputation models of quantitative molecular traits, and each tool has its own advantage for modeling a specific type of genetic architecture underlying the molecular trait of interest. For example, PrediXcan^11^ uses a penalized linear regression model with Elastic-Net penalty^12^ which has the advantage of modeling sparse genetic architectures. TIGAR^2^ implements the nonparametric Bayesian latent Dirichlet process regression (DPR) which has been shown effective to model the infinitesimal genetic architecture^13^. FUSION^5^ uses penalized linear regression model with Elastic-Net penalty and LASSO penalty^14^, regular linear regression model with best unbiased linear predictor (BLUP), single variant model with Top eQTL or pQTL (Top1), and Bayesian spare linear mixed model (BSLMM)^15^, and then selects the best model for gene-based association study based on cross-validation (CV) R^2^ of these multiple models.

Even though the FUSION tool employs multiple statistical models, it only uses the model with top performance based on reference data. Rather than selecting a single statistical method to estimate molecular QTL weights, previous study^16^ have shown that combining association test p-values based on multiple statistical methods using the Aggregated Cauchy Association Test^17^ (ACAT) can increase power while controlling for type I error. Therefore, we apply ACAT to aggregate PWAS p-values obtained by TIGAR, PrediXcan, and FUSION, which subsequently obtain an omnibus PWAS (PWAS-O) p-value.

PWAS-O is expected to improve PWAS power compared to PWAS based on a single imputation model of protein abundances, as PWAS-O better accounts for the unknown genetic architecture of protein abundances with multiple statistical models.

To evaluate the performance of PWAS-O, we conducted simulation studies using real genetic data based on patterns observed in real PWAS. We showed that PWAS-O outperformed all three individual methods with well calibrated type I error. We then applied PWAS-O to study AD dementia by using the reference human proteome-wide data of dorsolateral prefrontal cortex (DLPFC) of 355 older decedents with whole genome sequencing (WGS) genetic data, and summary data from the most recent GWAS of AD dementia (n=762,917, excluding 23&me samples)^1^. Next, we examined the protein-protein interaction (PPI) network among the identified PWAS risk genes of AD dementia. We identified 43 risk genes of AD dementia by PWAS-O that were connected through a protein-protein interaction (PPI) network involving the well-known risk genes of AD dementia. We validated the potential causality of 27 out of 43 (63%) PWAS risk genes by probabilistic Mendelian randomization (PMR-Egger)^18^ and summary data-based Mendelian randomization (SMR)^19, 20^ tool.

## Results

### Omnibus PWAS (PWAS-O)

Similar to the standard two-stage PWAS/TWAS method (**Methods; Fig. 1**)^11, 21^, protein abundance imputation models are first trained in Stage I to obtain estimated effect sizes of genetic variants/predictors (i.e., pQTL weights), by TIGAR^21^ (nonparametric Bayesian DPR), PrediXcan^11^ (Elastic-Net), and FUSION^5^ (Elastic-Net, LASSO, BLUP, Top1). Genetic variants within ±1MB around the transcription starting and termination sites of the corresponding coding gene were considered as predictors in the imputation models. In Stage II, these trained pQTL weights (coefficients of these genetic variant predictors) are used as variant weights along with GWAS data to conduct gene-based association tests. For each protein-coding gene, PWAS p-values will be respectively obtained by using the pQTL weights estimated by TIGAR, PrediXcan, and FUSION, which will then be combined by ACAT to generate the corresponding PWAS-O p-value (**Methods**). We present the PWAS-O framework in **Fig. 2**. Both individual-level and summary-level GWAS data can be used in Stage II. A reference panel of the same ancestry as the GWAS data can be used to provide LD information for gene-based association tests in Stage II. The TIGAR^21^ tool provides functions to calculate LD covariance matrices and conduct gene-based association tests.

**Fig. 1:**
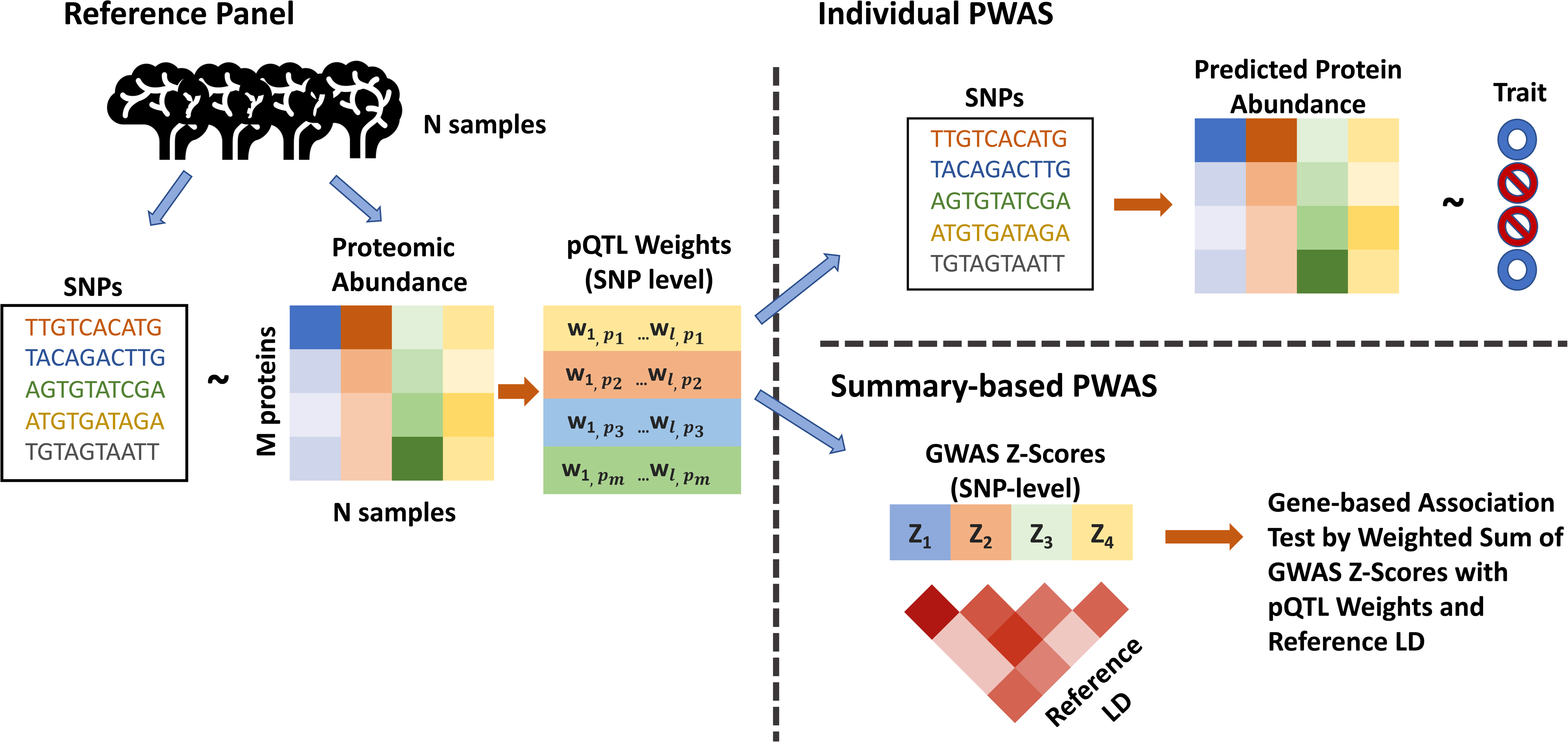
Two-stage PWAS framework. Stage I: Protein abundance imputation models are trained using *cis*-SNPs as predictors and proteomic abundance as the outcome, which estimates pQTL effect sizes (weights) per protein-coding gene. Stage II: Individual PWAS predicts the protein abundances using the trained imputation models and individual-level genotype data of test samples, and then tests the association between the predicted protein abundance and the trait of interest. Summary-based PWAS takes the pQTL effect sizes as variant weights to conduct an equivalent gene-based association test with GWAS Z-scores and reference LD.

**Fig. 2:**
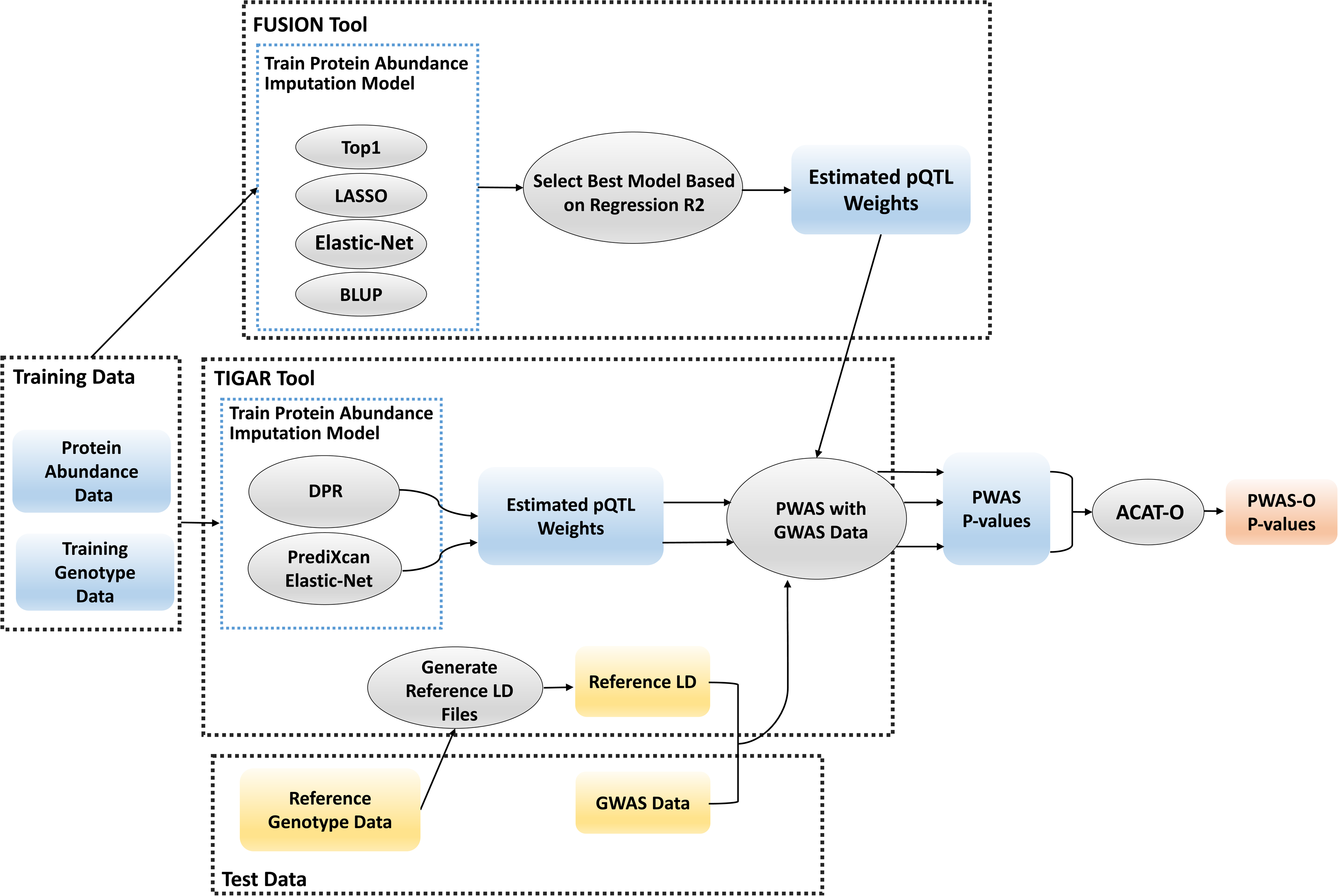
PWAS-O framework. First, train protein abundance imputation models using reference data and conduct two-stage PWAS (**Fig. 1**) using GWAS data by TIGAR, PrediXcan, and FUSION tools. Second, obtain a PWAS-O p-value per protein-coding gene by aggregating all PWAS p-values by ACAT.

### Simulation Study

Simulation studies were conducted to evaluate the performance of PWAS-O method. We use the real genotype data from 1894 whole genome sequencing (WGS) samples from the Religious Orders Study/Memory and Aging Project (ROS/MAP)^22^ and Mount Sinai Brain Bank (MSBB)^23^ study to simulate protein abundance data. We randomly selected 500 protein-coding genes with various gene length, and split samples into 568 training (30%) and 1326 testing (70%) samples to mimic a sample size in real reference panels for training protein abundance imputation models. We considered 4 different scenarios consisting of 2 proportions of causal cis-pQTL, *p_causal_* = (0.001, 0.01), and 2 proportions of protein abundance variance attributed to causal 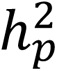 = (0.01, 0.05).

We generated protein abundance (*E_p_*) from the multivariable regression model 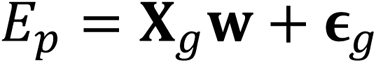, using genotype matrix **X**_g_ of the randomly selected causal pQTL of protein 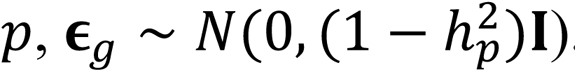. For each protein-coding gene, we generated 10 replicates of pQTL effect sizes **w** from *N*(0,1) per scenario, and rescaled these effect sizes to ensure the protein expression variance explained by causal variants is 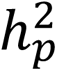. For each scenario, a total of 5000 replicates of protein traits were generated for selected 500 protein-coding genes. We then generated GWAS Z-scores from a multivariate normal distribution with 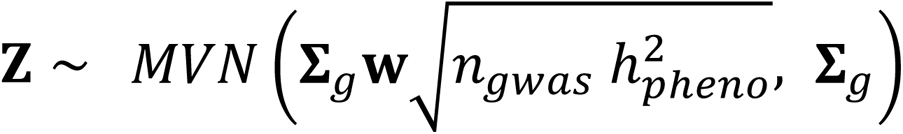^24^, where **w** denotes the true causal pQTL effect sizes, **Ʃ***_g_* is the correlation matrix of genotype **X***_test_*. for testing samples, and *n_gwas_* denotes GWAS sample size. We set 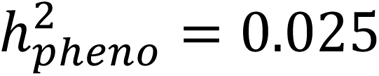, denoting the amount of phenotypic variance explained by simulated genetically regulated protein abundance **X***_g_***w**. We considered *n_gwas_* = (25K, 50K, 75K, 100K) for scenarios with 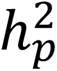 = 0.05 and *n_gwas_* = (600K, 800K, 1000K, 1200K) with 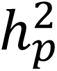 = 0.01. We generated 10 replicates of GWAS Z-scores per simulated protein trait per gene, resulting in a total of 50,000 PWAS simulation replicates.

In stage I of PWAS simulation analysis, we applied TIGAR, PrediXcan and FUSION to estimate pQTL weights from simulated protein traits of 568 training samples. In stage II, we integrated estimated pQTL weights with simulated GWAS Z-scores to conduct gene-based association tests. We further combined PWAS p-values obtained by these 3 tools using ACAT to generate PWAS-O p-values. We evaluated the training performance of these 3 individual methods by calculating the squared Pearson correlation coefficient between imputed genetically regulated protein abundances and simulated protein traits of 1326 testing samples, which was referred as test *R*^2^. PWAS power was given by the proportion of 50,000 repeated simulations with PWAS p-value value < 2.5 × 10^01^ (genome-wide significance threshold for gene-based association test). We compared PWAS power by PWAS-O to all 3 individual methods.

We showed that optimal test *R*^2^ and PWAS power based on one statistical model (**Fig. 3**) depended on the underlying genetic architecture of protein abundance (*p_causal_*) and protein heritability (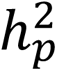). When protein heritability was small (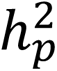 = 0.01), PrediXcan had the lowest test *R*^2^ and power. When protein heritability increased (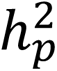 = 0.05) with sparse true cis-pQTLs (*p_causal_* = 0.001), PrediXcan became the optimal method with highest test *R*^2^ and power. For a less sparse model with *p_causal_*= 0.01, FUSION generally performed best among the individual methods. We found that PWAS-O obtained the highest power across all the scenarios by combining the PWAS p-values based on 3 methods.

**Fig. 3:**
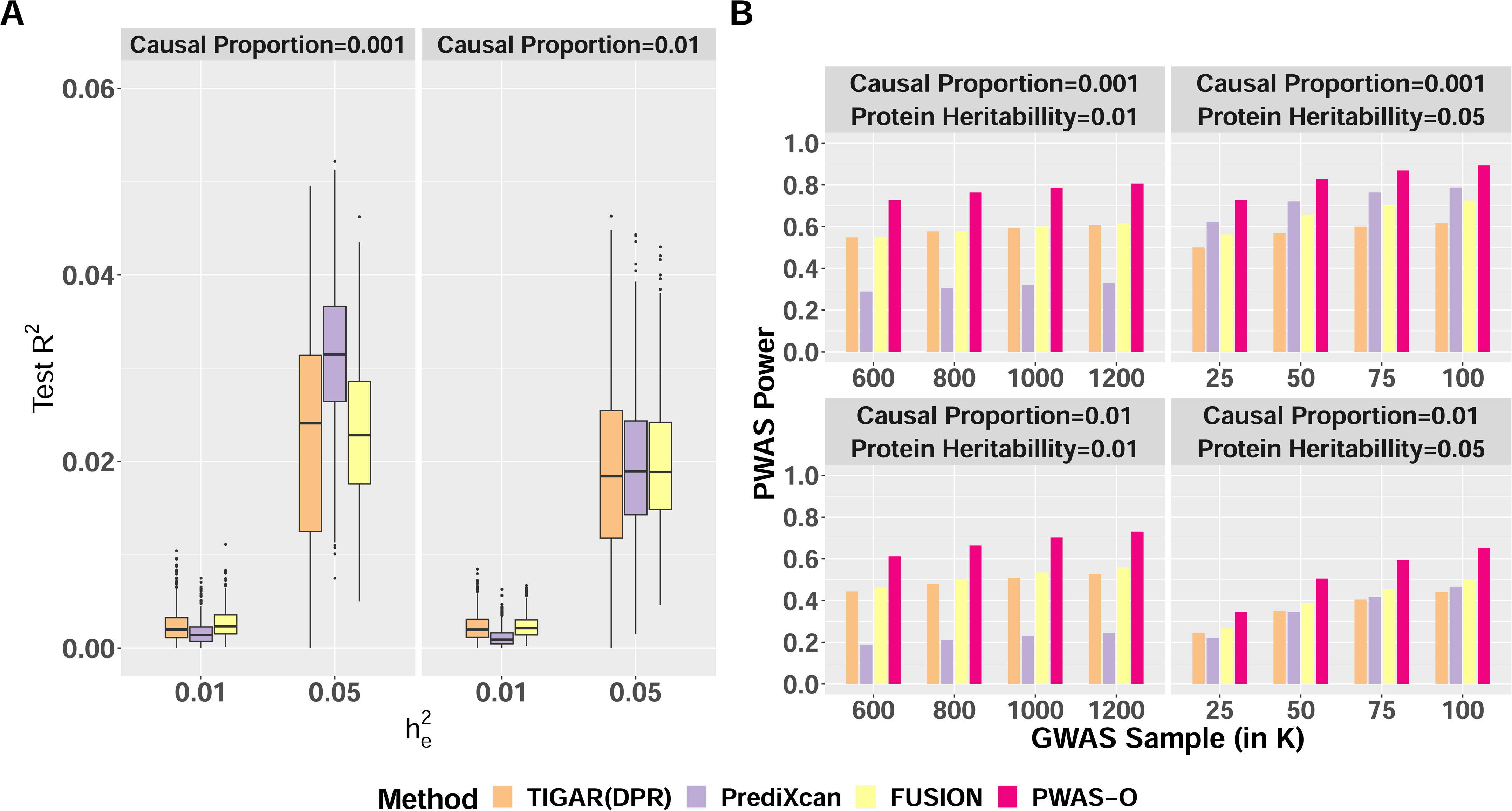
Test R^2^ (A) and PWAS power (B) comparison in simulation studies. Different scenarios with proportions of true causal cis-pQTL *p*_causal_ = (0.001,0.01) and protein expression heritability 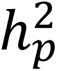 = (0.01, 0.05) were considered in the simulation studies. Boxplots of test R^2^ in 50,000 simulations per scenario for 3 PWAS tools were presented (A). Any simulation that was unsuccessful in training protein imputation models would contribute a zero value to the test R^2^ score. The median was depicted as a black bar, while the lower and upper hinges represented the 25th and 75th percentiles, respectively. Additionally, the whiskers extended from the hinges to a maximum of 1.5 times the interquartile range. Any data points beyond the whiskers were plotted individually. The power of 50,000 simulations were plotted with respect to various GWAS sample sizes, comparing PWAS-O to 3 PWAS tools (B).

We also evaluated type I errors of these individual methods and PWAS-O under one simulated scenario with 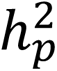 = 0.05 and *p_causal_* = 0.001. We simulated 2 × 10^2^ phenotype from *N*(0,1) for each of the 500 genes and permuted estimated pQTL weights to conduct a total of 10^6^ null simulations. Both individual methods and PWAS-O showed well calibrated type I errors, as shown by quantile-quantile (Q-Q) plots of p-values (**Fig. S1**).

### ROS/MAP Proteomics and WGS data

The human brain proteomic reference data used in the PWAS-O application study of AD dementia in this study were profiled from the DLPFC of postmortem brain samples donated by 400 participants of European ancestry of the Religious Orders Study and Memory and Aging Project (ROS/MAP)^7, 22^. These human proteome abundance data underwent quality control (**Methods; Fig. S2**) that regressed out the effects of technical factors and confounding covariates including age at death, sex, postmortem interval, and study (ROS or MAP). Proteome abundance data of 8,430 proteins of 355 individuals who also had WGS data profiled^25^ were obtained after quality control and used as reference data in the PWAS-O application study of AD dementia in this study. This ROS/MAP reference proteomic data set is one of the largest available proteomic data sets for DLPFC tissue which is a relevant tissue for studying AD dementia.

### PWAS-O results of studying AD dementia

At training Stage I, five-fold cross-validation was conducted to evaluate the training performance per statistical model. The protein abundance imputation models with 5-fold CV *R*^2^ >0.5% were retained and their estimated pQTL weights were used to conduct PWAS. As suggested by the TIGAR tool^21^, the threshold of 5-fold CV *R*^2^ >0.5% was arbitrarily chosen to allow a greater number of protein-coding genes to be tested by the follow-up gene-based association study (i.e., PWAS). The threshold of CV *R*^2^ would not affect the statistical validity of the test statistic used in the follow-up PWAS.

As shown in **Table 1**, 6,673 protein abundance imputation models were retained by TIGAR with median CV *R*^2^ = 0.013; 1,835 protein abundance imputation models were retained by PrediXcan with median CV *R*^2^ = 0.0094; and 2,173 protein abundance imputation models were retained by FUSION (the best model selected out of LASSO, Elastic-Net, BLUP, Top1 models with the highest CV *R*^2^) with median CV *R*^2^ = 0.0093. Histograms of the CV *R*^2^ obtained by these three tools were presented in **Fig. S3**.

**Table 1.** The total number of tested proteins with CV *R*^2^ > 0.005 for the corresponding protein abundance imputation models trained using ROS/MAP reference data by TIGAR, PrediXcan, and FUSION tools. Median CV *R*^2^ of tested proteins are presented in the third column.

| Model | Number of Proteins | Median $CV R^2$ |
| --- | --- | --- |
| TIGAR | 6,673 | 0.013 |
| PrediXcan | 1,835 | 0.0094 |
| FUSION | 2,173 | 0.0093 |

As described in the above about the PWAS-O framework, we first conducted PWAS by employing TIGAR^21^, PrediXcan^11^, and FUSION^5^ tools, using the ROS/MAP reference proteomics data of DLPFC and the most recent GWAS^1^ summary data of AD dementia (n=762,917, excluding 23&me samples; **Methods**). That is, the estimated pQTL effect sizes in the retained protein abundance imputation models by these three tools were taken as variant weights to conduct gene-based association tests along with GWAS summary data of AD dementia (**Fig. S4**). Second, we implemented ACAT^17^ to obtain PWAS-O p-values from the PWAS p-values by these three tools. Q-Q plots of PWAS-O and PWAS results of AD dementia by these three tools were presented in **Fig. S5**.

Our PWAS-O approach identified 43 significant genes with false discovery rate (FDR) (i.e., q-value) <0.05, which could affect the risk of AD dementia through their genetic effects on the corresponding protein abundances (**Fig. 4**). We curated 19 independent genes from these 43 significant genes (**Table 2; Table S1**), by considering only the most significant genes among all significant genes with shared test regions (±1MB around the test transcription starting and termination sites of the protein-coding gene). In particular, we found all individual methods contributed to the PWAS-O results (**Table 2; Table S1; Fig. S6**). Out of 43 PWAS risk genes, 33, 9, and 7 genes were detected by TIGAR, FUSION, and PrediXcan, respectively.

**Table 2.** Independent PWAS-O risk genes of AD dementia. Missing P-values denoted by dashes are due to either CV *R*^2^ < 0.5% for the corresponding protein abundance imputation models or no trained protein abundance imputation model by PrediXcan due to variable selection.

| Gene | CHR | PWAS-O<br>P-value | TIGAR<br>P-value | PrediXcan<br>P-value | FUSION<br>P-value |
| --- | --- | --- | --- | --- | --- |
| CDC42<sup>c</sup> | 1 | $2.47 \times 10^{-4}$ | $2.47 \times 10^{-4}$ | - | - |
| ERCC3<sup>b</sup> | 2 | $2.91 \times 10^{-4}$ | $1.80 \times 10^{-4}$ | - | $7.54 \times 10^{-4}$ |
| MGAT5 | 2 | $6.38 \times 10^{-5}$ | $7.62 \times 10^{-4}$ | $2.19 \times 10^{-5}$ | $2.97 \times 10^{-2}$ |
| HSPA1B<sup>b</sup> | 6 | $2.64 \times 10^{-9}$ | - | - | $2.64 \times 10^{-9}$ |
| TULP1<sup>b</sup> | 6 | $2.88 \times 10^{-4}$ | $4.14 \times 10^{-1}$ | - | $1.44 \times 10^{-4}$ |
| PLA2G7<sup>c</sup> | 6 | $9.03 \times 10^{-5}$ | $9.03 \times 10^{-5}$ | - | - |
| MAP3K7 | 6 | $5.71 \times 10^{-5}$ | $5.71 \times 10^{-5}$ | - | - |
| TRIM4<sup>b</sup> | 7 | $3.72 \times 10^{-5}$ | $3.72 \times 10^{-5}$ | - | - |
| AGFG2<sup>a,b,c</sup> | 7 | $9.86 \times 10^{-8}$ | $1.24 \times 10^{-7}$ | - | $8.20 \times 10^{-8}$ |
| ZYX<sup>b,c</sup> | 7 | $1.39 \times 10^{-5}$ | $7.91 \times 10^{-6}$ | $5.86 \times 10^{-5}$ | - |
| C1QTNF4<sup>b,c</sup> | 11 | $2.24 \times 10^{-5}$ | $2.24 \times 10^{-5}$ | - | - |
| MRPL16<sup>b,c</sup> | 11 | $8.66 \times 10^{-15}$ | $4.33 \times 10^{-15}$ | $1.31 \times 10^{-2}$ | - |
| CCDC86<sup>b</sup> | 11 | $2.83 \times 10^{-9}$ | $1.42 \times 10^{-9}$ | - | $1.59 \times 10^{-1}$ |
| DIABLO | 12 | $9.26 \times 10^{-5}$ | $9.26 \times 10^{-5}$ | - | - |
| ATPAF2<sup>b</sup> | 17 | $8.20 \times 10^{-5}$ | $8.20 \times 10^{-5}$ | - | - |
| DCAKD<sup>b</sup> | 17 | $6.19 \times 10^{-5}$ | $3.62 \times 10^{-3}$ | - | $3.12 \times 10^{-5}$ |
| CLPTM1<sup>a,b,c</sup> | 19 | $1.44 \times 10^{-70}$ | $1.49 \times 10^{-21}$ | $7.21 \times 10^{-71}$ | - |
| SPHK2<sup>c</sup> | 19 | $1.76 \times 10^{-4}$ | $4.87 \times 10^{-2}$ | $8.82 \times 10^{-5}$ | - |
| YWHAB | 20 | $1.71 \times 10^{-4}$ | $1.71 \times 10^{-4}$ | - | - |

<sup>a</sup> Known GWAS risk genes of AD dementia.
<sup>b</sup> Known TWAS risk genes of AD dementia, or test region overlapped with TWAS risk genes.
<sup>c</sup> Significant mediated causal genetic effects by PMR-Egger or SMR.

**Fig. 4:**
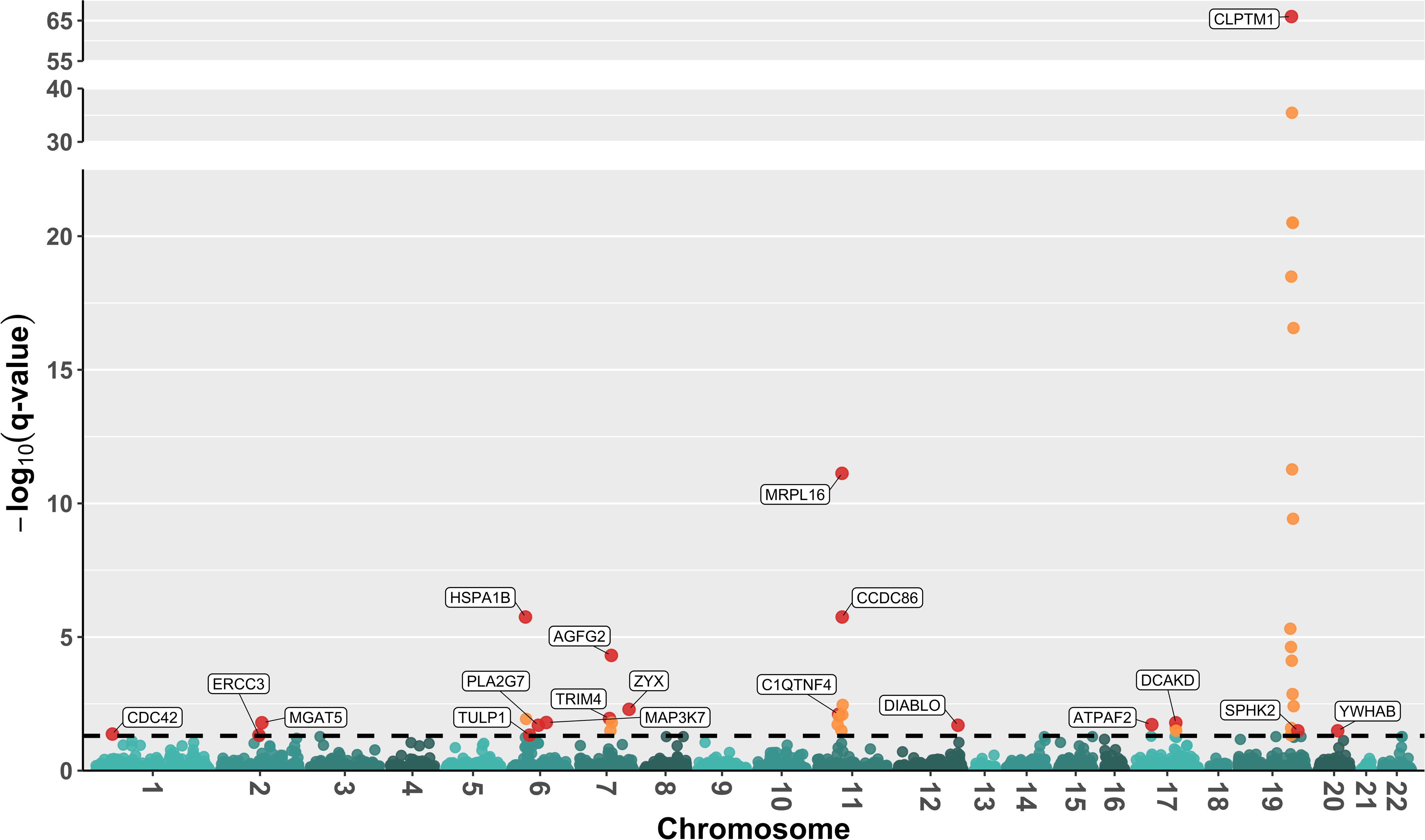
M**a**nhattan **plot of PWAS-O results for AD dementia.** A total of 43 significant risk genes were identified with FDR q-value < 0.05. A total of 19 independently significant genes were labeled. The –log10(q-values) were plotted in y-axis, and –log10(0.05) was plotted as the dashed horizontal line.

We found 10 out of these 43 PWAS risk genes were detected by previous GWAS studies of AD as shown in GWAS Catalogue^26^, including *AGFG2*^27^*, CLPTM1*^28^*, PVR*^28^*, PVRL2*^28^*, APOC1*^29^*, APOC2*^30^*, CKM*^28^*, ERCC1*^31^*, OPA3*^31^, and *SYMPK*^31^. PWAS risk genes *CDC42*^32^, *MAP3K7*^33^, and *FNBP4*^32^ were found associated with cognitive ability in previous GWAS studies. PWAS risk genes *CCDC86*^34^*, KIF18B*^34^, *PPM1N*^34^, and *RTN2*^34^ were previously found associated with neurofibrillary tangles measurement (a key clinical indicator of AD) by GWAS studies. Additionally, 13 out of 43 significant AD PWAS genes (*ZYX*^35^*, C1QTNF4*^36^*, FNBP4*^35^*, PVR*^3^*, PVRL2*^36^*, TOMM40*^3^*, APOC1*^4^*, APOC2*^37^*, CLPTM1*^36^*, ERCC1*^38^*, OPA3*^38^*, EML2*^4^ and *DMWD*^35^) were significant risk genes of AD by previously published TWAS.

Our PWAS-O findings not only replicated significant risk genes identified by previous GWAS and TWAS, but also identify novel risk genes for AD dementia. PWAS-O identified 5 novel risk genes that were neither detected by GWAS nor TWAS (*MGAT5, PACSIN3*, *PLA2G7, DIABLO, YWHAB*). The novel gene *PLA2G7* was shown to have biological function involved in inflammatory and oxidative activities^39^, with a known association with AD and cognitive decline^40^. The novel gene *PACSIN3* has been implicated in the disorders of the central nervous system such as schizophrenia^41^.

### pQTL weights by different regression models

To investigate the genetic architecture underlying protein abundances, we plotted the pQTL weights estimated by TIGAR (DPR), PrediXcan (Elastic-Net), and FUSION (best model out of LASSO, Elastic-Net, BLUP, Top1) for significant PWAS risk genes *PPM1N* and *DCAKD* in **Fig. 5**. Generally, TIGAR and FUSION had non-zero pQTL weights estimated for almost all test SNPs within the test region, for using a model assuming infinitesimal genetic architecture^13^. Whereas, PrediXcan had much fewer non-zero pQTL weights estimated for assuming a sparse genetic architecture and enforcing variable selection. For example, *PPM1N* was a significant risk gene identified by both TIGAR and FUSION (**Table 2**), which has hundreds of test SNPs with non-zero pQTL weights (by both TIGAR and FUSION) that were colocalized with significant GWAS signals (colored dots in **Fig. 5A**). Gene *DCAKD* was only found significant by FUSION (p-value = 3.12 × 10^03^) but not by TIGAR (p-value = 3.62 × 10^02^), for having fewer test SNPs of relatively large magnitude of pQTL weights that were colocalized with significant GWAS signals (**Table2,** **Fig. 5B**).

**Fig. 5:**
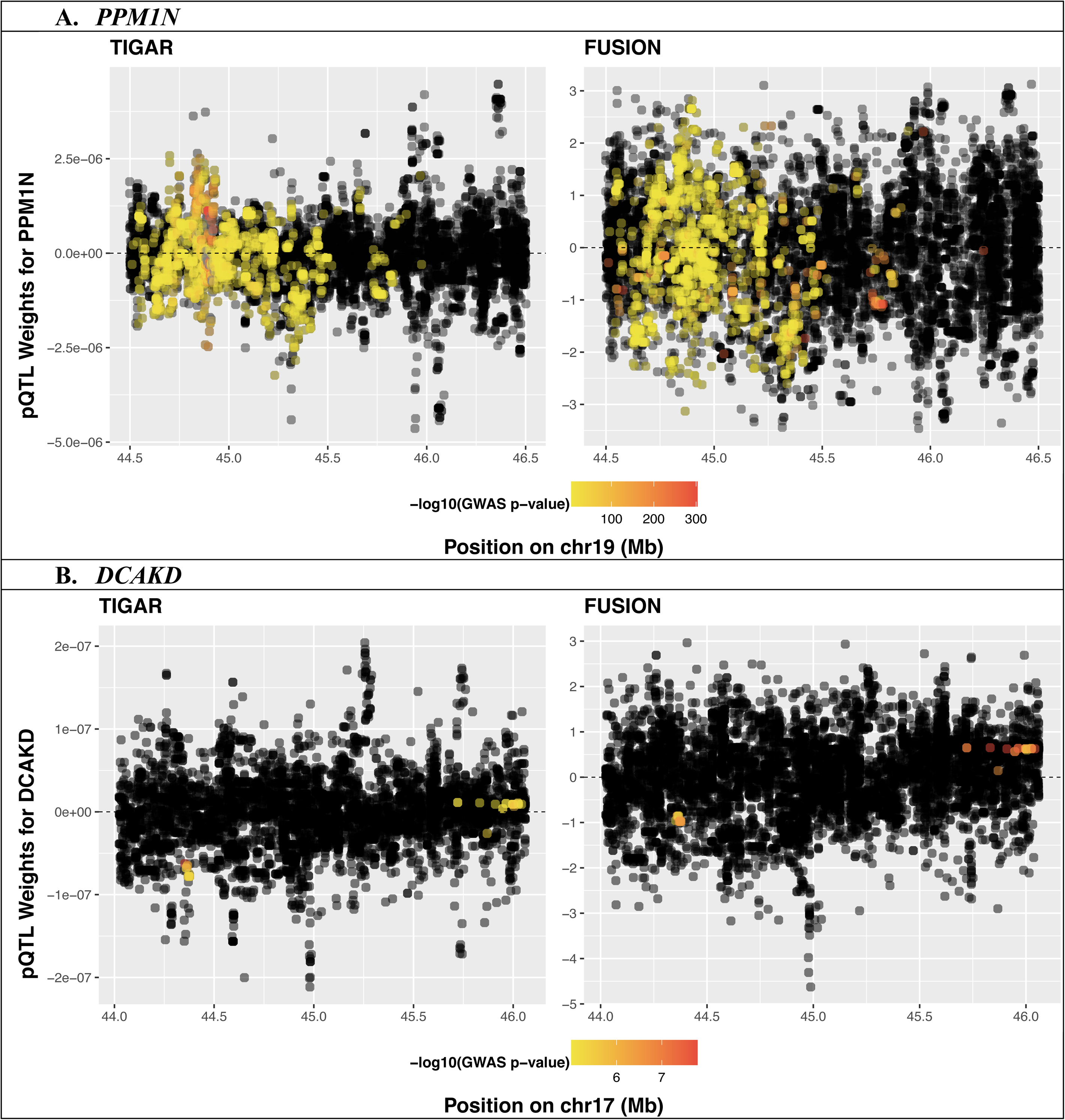
pQTL weights estimated by TIGAR (DPR) and FUSION (best model of LASSO, Elastic-Net, BLUP, Top1) for example significant genes *PPM1N* (A) and *DCAKD* (B) by PWAS-O. The pQTL weights were plotted in the y-axis for all test SNPs in the test gene region, with color coded with respect to their –log10 (GWAS p-value). SNPs with GWAS p-value < 10^-3^ were plotted in yellow color. Gene *PPM1N* (A) was found significant by both TIGAR and FUSION, as multiple test SNPs with non-zero pQTL weights (by TIGAR and FUSION) were colocalized with significant GWAS SNPs (yellow dots). Whereas, the significance of *DCAKD* (B) was mainly driven by FUSION, for having fewer SNPs with significant GWAS p-values that are also colocalized with a relatively large magnitude of pQTL weights.

Similarly, comparing pQTL weights obtained by TIGAR to PrediXcan for significant PWAS genes *MBLAC1* and *MRPL16* (**Fig. S7**), we observed that less than 10 test SNPs had non-zero pQTL weights by PrediXcan, versus thousands of test SNPs with non-zero pQTL weights by TIGAR. Because PrediXcan estimated zero pQTL weights for most SNPs with significant GWAS p-values in the test region of gene *MRPL16* (**Table 2**, **Fig. S7A**), *MRPL16* was only found significant by TIGAR (p-value = 4.33 × 10^0^^43^), but not by PrediXcan (p-value = 1.31 × 10^0’^). Whereas, when both TIGAR and PrediXcan estimated non-zero pQTL weights for SNPs with significant GWAS p-values in the test region of gene *MBLAC1* (**Table S1, Fig. S7B**), it was identified significant by both TIGAR (p-value = 7.31 × 10^03^) and PrediXcan (p-value = 3.49 × 10^05^).

These scatterplots of pQTL weights of PWAS risk genes showed that significant PWAS genes were mainly driven by test SNPs that had non-zero pQTL weights colocalized with significant GWAS signals. Because the underlying genetic architectures of protein abundances are complicated and unknown in practice, no single statistical model would lead to superior PWAS power across genome-wide protein-coding genes. Our proposed PWAS-O approach is expected to gain power in practice for considering multiple statistical models per protein.

### PPI network among PWAS risk genes of AD dementia

Next, we used STRING^42, 43^ to investigate the PPI network among these 43 PWAS risk genes of AD dementia, by using public data sources of known protein-protein interactions including evidences from curated databases, experiments, text mining, co-expression, gene fusions, and protein homology^42^ (**Methods**). STRING clustered 23 out of these 43 PWAS risk genes into 4 communities (**Fig. 6**), where different data sources of the protein-protein connection were depicted by differently colored connection edges. It is interesting to see that the largest community of these PWAS risk genes is a network involving genes *TOMM40*, *APOC1*, and *APOC2* that are located nearby the well-known risk gene *APOE* of AD dementia, and are known to be associated with both AD dementia and low-density lipoprotein levels^44^.

**Fig. 6:**
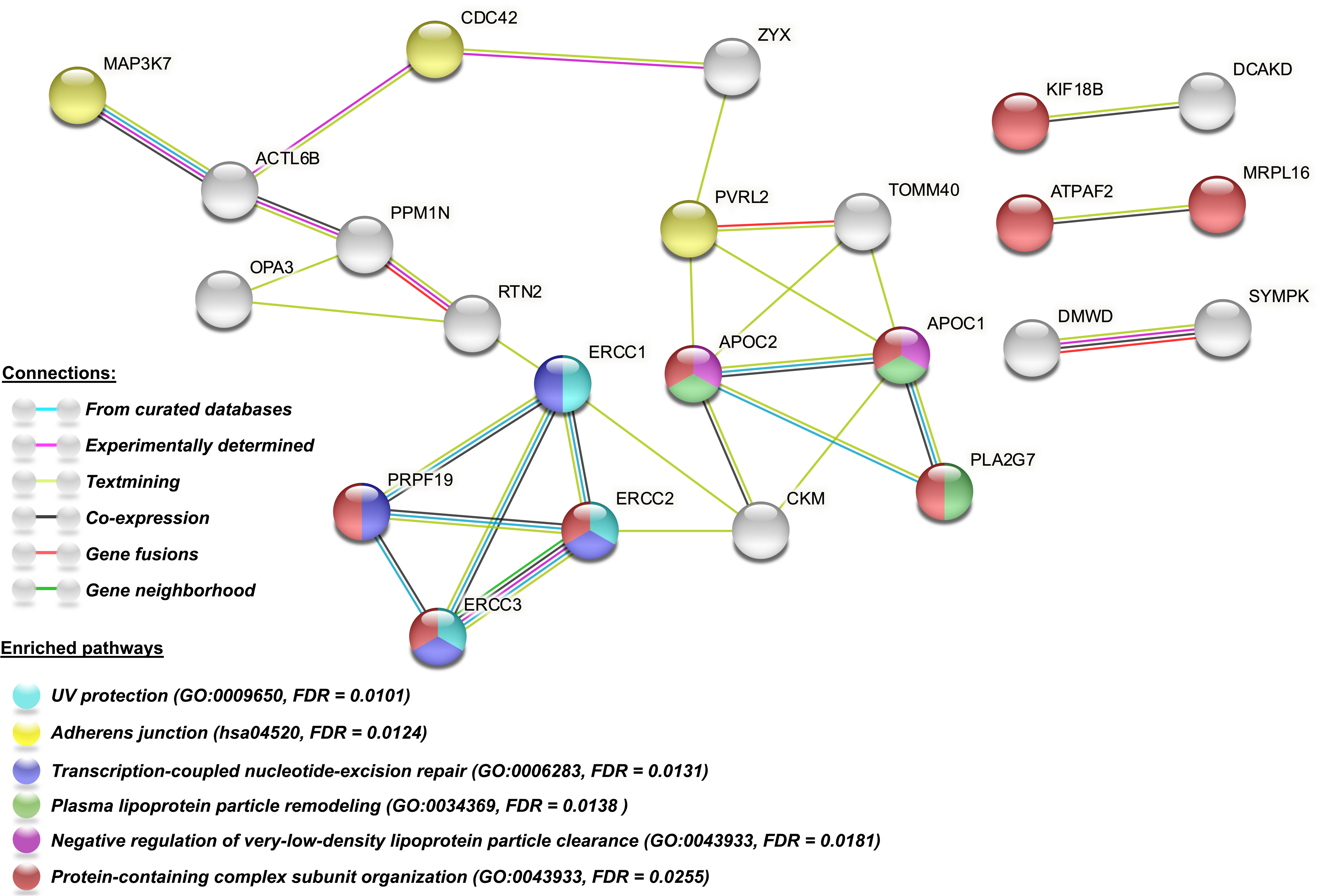
P**P**I **networks and enriched pathways among 43 PWAS risk genes of AD dementia by STRING.** Edges represent physical PPI, with different colors representing different sources of connection evidence. Node colors represent different enriched GO terms or KEGG pathways with FDR < 0.05.

STRING also provided gene enrichment analysis results with respect to Gene Ontologies (GO) and KEGG pathways. We observed that these 43 PWAS risk genes were enriched for the GO terms of UV protection (*ERCC1, ERCC2, ERCC3*) with FDR = 0.010, transcription-coupled nucleotide-excision repair (*ERCC1, ERCC2, ERCC3, PRPF19*) with FDR = 0.013, plasma lipoprotein particle remodeling (*APOC1, APOC2, PLA2G7*) with FDR = 0.014, negative regulation of very-low-density lipoprotein particle clearance (*APOC1, APOC2*) with FDR = 0.018, protein-containing complex subunit (*MRPL16, ATPAF2, KIF18B, ERCC2, ERCC3, PRPF19, APOC1, APOC2, PLA2G7*) with FDR = 0.026, and the KEGG pathway of adherens junction (*MAP3K7*, *CDC42*, *PVRL2*) with FDR = 0.012.

Interestingly, novel PWAS risk genes *ERCC3*, *ERCC2* and *PRPF19* are closely interconnected with a known GWAS/TWAS risk gene *ERCC1* in the PPI network and enriched in nucleotide-excision repair pathway. Novel PWAS risk gene *PLA2G7* connects with *APOC1* and *APOC2* in the PPI network and involves in the plasma lipoprotein particle remodeling pathway.

These PPI network analysis results showed that the list of identified risk genes by PWAS-O were connected with known risk genes of AD dementia and enriched in AD related pathways.

### Examine causal and pleiotropy effects of PWAS risk genes

We applied both probabilistic Mendelian randomization (PMR-Egger)^18^ and summary data-based Mendelian randomization (SMR) methods to test if the causal genetic effects on AD dementia were mediated through protein abundance in the possible presence of horizontal pleiotropy (**Methods**).

Horizontal pleiotropy means that genetic effects on AD dementia and on protein abundances originated from the same SNPs but through different pathways (i.e., with no mediation effects). PMR-Egger suggested that 25 out of 43 PWAS risk genes have significant causal effects (FDR<0.05) on AD dementia after controlling for potential horizontal pleiotropy effects, including 6 genes (*PVRL2*, *TOMM40, APOC1, RTN2, OPA3, EML2*) nearby the *APOE* region that also have significant horizontal pleiotropy effects with FDR<0.05. Similarly, the method of SMR with its accompanying heterogeneity in dependent instruments (HEIDI) test found that 22 out of 43 PWAS risk genes have potential causal effects on AD dementia with FDR<0.1, of which 20 genes were overlapped with 25 causal genes detected by PMR tool. These results validated potential causal genetic effects that were mediated through protein abundances for 27 (63%) PWAS risk genes identified by PWAS-O (**Table 2; Table S1**).

### Compare with TWAS results of AD dementia

To determine whether the 43 PWAS risk genes had similar evidence at the transcript level, we also conducted TWAS of AD dementia using ROS/MAP transcriptomic reference data of DLPFC tissue and the same GWAS summary data of AD dementia by TIGAR and PredXican. From TWAS p-values by these two tools, we obtained omnibus TWAS (TWAS-O) p-values by ACAT (**Methods**). We found that 34 out of 43 PWAS risk genes (79%) were either detected by TWAS-O or located within 1MB of the transcription starting sites and termination sites of the significant risk genes by TWAS-O with FDR<0.05 (**Table S2, Fig. S8**). These results showed that the majority of PWAS risk genes had genetic effects on phenotype that were mediated through both transcriptome and proteome.

## Discussion

In this work, we propose an omnibus PWAS approach (PWAS-O) that improves power for better modeling of the unknown genetic architecture of proteome abundances by multiple statistical models. By aggregating PWAS p-values obtained from several available tools (i.e., TIGAR, PrediXcan, and FUSION) that used different statistical models, our PWAS-O approach improves power over using one individual tool. Our simulation studies showed that the performance of each of these three tools depended on the underlying genetic architecture for protein abundance and protein heritability. We found PrediXcan performed better for sparse architecture and higher heritability, whereas TIGAR and FUSION had better performance for denser architecture. PWAS-O was demonstrated with optimal performance across all considered simulation scenarios with well calibrated type I error. Since the genetic architecture of protein abundance is unknown, the practical use of PWAS-O is well justified.

By using the ROS/MAP reference proteomic data and the most recent GWAS summary data of AD dementia, PWAS-O identified 43 risk genes of AD dementia. All three individual methods contributed to the PWAS-O results. Notable biological roles were shown by previous studies for these identified PWAS risk genes include lipoprotein metabolism, inflammatory reactions, DNA repair, pre-mRNA splicing, and mitochondrial function. For example, *APOC1* and *APOC2*, located in a gene cluster on chromosome 19 including the well-known AD risk gene *APOE*^45, 46^, play important roles in the development of lipoprotein metabolism^44^. Especially, *APOE* is an intensively studied risk gene of AD and plays an essential role in the central nervous system^47, 48^. Connected with *APOC1* and *APOC2,* as shown in the PPI of our identified PWAS risk genes, *PLA2G7* is a novel AD risk gene identified by PWAS-O and associated with inflammatory oxidative activities, and has been implicated in cognitive impairment and AD^39^.

Additionally, *ERCC1, ERCC2,* and *ERCC3* are involved in nucleotide excision repair pathway contributing to genome stability^49, 50^, which plays a critical role during the development of the central nervous system^51^ and whose impairment is associated with AD^52^. Gene *PRPF19* that is interconnected with *ERCC1, ERCC2*, and *ERCC3* encodes a pre-mRNA processing factor involved in regulating pre-mRNA splicing^53^. Prior studies indicated that *PRPF19* had a neuronal function and was implicated in neurological diseases^54, 55^; lower expression level of gene *MRPL16* encoding a mitochondrial ribosome protein was observed in AD blood cells which may reflect a general response to inflammatory environment in AD^56^; gene *ATPAF2* connected with *MRPL*16 encodes a protein involved in mitochondrial function and lipid pathway, which has been implicated in dementia pathogenesis^57^.

By STRING analysis, we showed that out of these 43 PWAS significant risk genes, 9 genes were enriched in the pathway of protein-containing complex subunit; 4 genes were enriched in the gene ontology pathway of transcription-coupled nucleotide-excision repair; 3 genes were enriched in the pathway of plasma lipoprotein particle remodeling. Compared to TWAS-O results using the same GWAS summary data, 34 out of 43 significant PWAS risk genes were either TWAS-O risk genes or within 1MB test region of TWAS risk genes. These results showed that PWAS risk genes identified by our PWAS-O approach are part of key biological pathways involving brain functions. The genes and proteins identified may be high-value therapeutic targets for further drug discovery as well as for mechanistic studies that can advance our understanding of the biology driving AD dementia in aging brains.

PWAS-O still has its limitations. First, the CV *R*^2^ is generally low for majority protein abundance prediction models. This might be due to small sample sizes in the reference proteomic data, or low heritability of protein abundances due to cis-SNPs. Only *cis*-SNPs within the ±1*MB* region around the transcription starting and termination sites of the corresponding protein-coding gene were considered as predictors for protein abundances by PWAS-O. Trans-SNPs could also be considered as additional predictors in the protein abundance prediction models, which might increase the CV *R*^2^ of protein abundance imputation models but will be subject to high computation burden^3^. Second, the standard two-stage PWAS method cannot distinguish whether causal SNPs for the phenotype of interest are only colocalized with pQTLs but have no mediation effect through the protein abundances (i.e., horizontal pleiotropy) or indeed have genetic effects mediated by protein abundances (i.e., vertical pleiotropy).

Horizontal pleiotropy happens when genetic variants impact the outcome through pathways different from or additional to the proteome abundances of the test gene^58^. Here, we applied PMR-Egger^18^ and SMR^59^ tool to test the causal effects of 43 PWAS risk genes while controlling for horizontal pleiotropy. We validated the potential causality 27 genes out of 43 genes identified by our PWAS-O. Third, a cluster of significant PWAS risk genes are likely to be identified as shown in the locus around *APOE* for AD dementia for sharing test SNPs in the overlapped test gene region. Even though the PMR-Egger and SMR tools provide some insights into the potential causality/mediation role of the genetically regulated protein abundances for the phenotype of interest, true causal genes will still need to be replicated by further biological experiments.

In summary, PWAS-O represents the first omnibus PWAS method that increases power by leveraging multiple statistical models. PWAS-O can be easily applied to integrating reference proteomic data with publicly available large-scale GWAS summary data, for studying complex polygenic diseases. We believe that PWAS-O would be useful for mapping risk genes with genetic effects potentially mediated through proteome abundances for complex diseases. Identified PWAS risk genes would shed light upon future studies of disease pathogenesis and therapeutics.

## Methods

### Two-stage PWAS

As shown in **Fig. 1**, the two-stage PWAS first trains protein abundance imputation models (Stage I) by taking *cis*-genotype data, ±1MB around the transcription starting and termination sites of the corresponding protein-coding gene, as predictors (X), protein abundance level (**E**_*p*_) as the outcome, as shown in the following linear regression model:

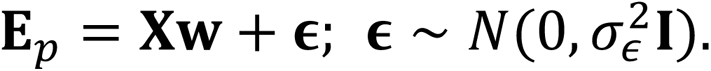

The effect size vector **w** could be estimated by different statistical models as employed by TIGAR^21^, PrediXcan^11^, and FUSION^5^, which could be viewed as a broad sense of pQTL effect sizes (also referred to as pQTL weights). Specifically, PrediXcan employs a penalized linear regression model with Elastic-Net penalty^11^; FUSION^5^ uses penalized linear regression model with Elastic-Net penalty and LASSO penalty^14^, regular linear regression model with best unbiased linear predictor (BLUP), single variant model with Top pQTL (Top1), and Bayesian spare linear mixed model (BSLMM)^15^, and then selects the best model according to CV *R*^2^; TIGAR^2^ implements the nonparametric Bayesian latent DPR model.

In Stage II, with individual-level GWAS data, the protein abundances will first be imputed by using the trained imputation models and individual-level genotype data of test samples. Then PWAS will test the association between the imputed protein abundances and the phenotype of interest. With summary GWAS data, PWAS would take the pQTL weights estimated in Stage I as variant weights to conduct gene-based association test with GWAS Z-scores and reference LD, which has been shown to be equivalent as testing the association between imputed genetically regulated protein abundances and the phenotype of interest^60^. That is, the PWAS test is equivalent to a burden test with the following test statistic:

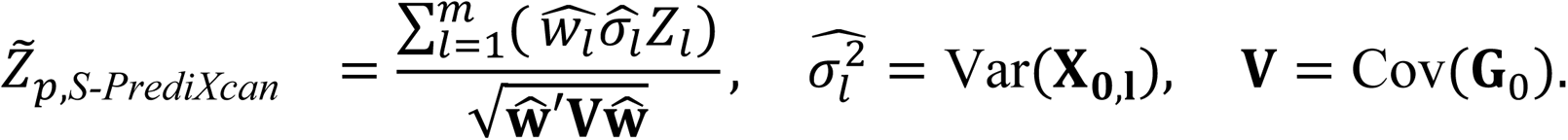

Here, 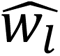 denotes the pQTL effect size estimates from protein imputation models; *Z_l_* denotes the Z-score statistics of single variant *l* by GWAS; and **V** denotes the LD covariance matrix from a reference panel of the same ancestry as the test GWAS cohort. The genotype variance 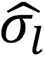 of test SNPs can also be estimated from a reference panel of the same ancestry as the test cohort.

### PWAS-O Method

As shown in **Fig. 2**, with PWAS p-values obtained by two-stage PWAS tools including TIGAR^21^, PrediXcan^11^, and FUSION^5^, PWAS-O uses ACAT^17^ to aggregate these three PWAS p-values to generate an omnibus test p-value per protein-coding gene. The ACAT test statistic is given by

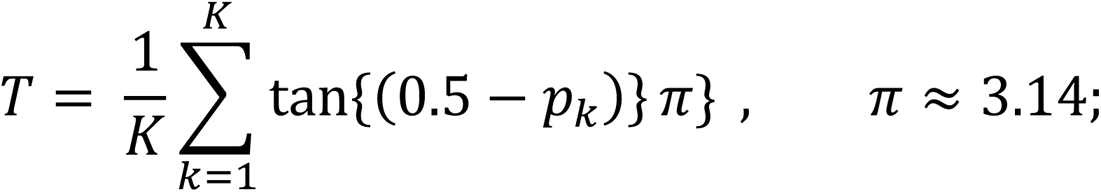

where *p_k_* is the PWAS p-value obtained by standard two-stage PWAS tools. Under null hypothesis, the p-value *p_k_* is uniformly distributed in [0, 1] and the transformation tan{(0.5 − *p_=_*)} is Cauchy distributed. Thus, the null distribution of the test statistic *T* can be approximated by a Cauchy distribution^18^ and the corresponding PWAS-O p-value can be approximated by

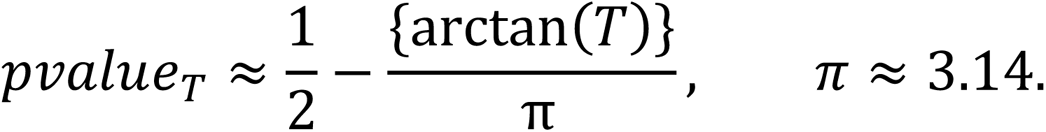

### ROS/MAP Proteomics and WGS Data

Human brain proteomic profiles were generated from the dorsolateral prefrontal cortex (DLPFC) of postmortem brain samples donated by 400 participants of European ancestry of the Religious Orders Study or Rush Memory and Aging Project (ROS/MAP)^7, 22^. Participants in ROS and MAP enroll without known dementia and agree to annual clinical evaluation and organ donation at death. All participants sign an informed consent, Anatomic Gift Act and Repository Consent. Both studies were reviewed by an Institutional Review Board of Rush University Medical Center. Age was calculated from birth date and date of death. Sex was self-reported. Protein measurements from DLPFC were quantified by tandem mass spectrometry.

As described in previous publication^61^, protein abundances were normalized by scaling each protein abundance with respect to a sample-specific total protein abundance, and proteins with missing values in more than 50% of the participants were excluded from the analyses. Protein abundance ratios related to baseline (sample-specific total protein abundance) were log_2_ transformed, with missing values imputed by the corresponding mean in the cohort. Confounding covariates of clinical characteristics including sex, age at death, postmortem interval, and study type (ROS or MAP), as well as technical factors including sequencing batch and MS2 versus MS3 mass spectrometry reporter quantification mode, were regressed out from protein abundances. Outlier samples were removed using iterative principal component analysis (PCA) as shown in **Fig. S2**. That is, samples with one of the top 2 principal components (PCs) greater than the corresponding four times of the standard deviations from the mean of the corresponding PC^61^.

The genotype data of ROS/MAP samples were profiled by whole genome sequencing (WGS)^25^. SNPs with Hardy-Weinberg equilibrium p-values <10^03^ and minor allele frequency >1% were used as genetic predictors for training the protein abundance imputation models. After quality control of both proteomic and genetic data, a total of 355 samples with both proteomic and WGS data were used as the reference data for the application study of PWAS-O in this study.

### Summary-level GWAS data of AD dementia

The summary-level GWAS data of AD dementia (i.e., single variant Z-score test statistics obtained by meta-analysis) were generated by the latest GWAS by Wightman et al^1^, including 762,917 individuals from 12 cohorts except 23&Me samples. About 11.3% of participants had clinically diagnosed AD dementia.

### PWAS of AD dementia

In this study, we used the ROS/MAP proteomics and WGS data of 355 samples as reference data to train protein abundance imputation models. For each protein, we first used TIGAR tool to train nonparametric Bayesian DPR model and Elastic-Net regression model (as used by PrediXcan), and then used FUSION^5^ tool to train LASSO, Elastic-Net, BLUP, and Top1 models and then selected the best model according to the CV *R*^2^. We did not train a BSLMM model due to its high computation cost for WGS data, and we expect that the nonparametric Bayesian DPR model would account for the mixed sparse and infinitesimal genetic architecture. The 5-fold CV was conducted for all protein abundance imputation models. The average CV *R*^2^ across 5 folds of validation data per protein was used to select the best performing FUSION model as well as filter protein abundance imputation models for the follow-up PWAS.

Second, protein abundance imputation models with 5-fold CV *R*^2^ >0.005 were considered as valid, and the corresponding pQTL weights were used along with summary-level GWAS data of AD dementia to test for PWAS risk genes^21^. Last, PWAS-O p-values were generated by aggregating PWAS p-values obtained by using pQTL weights estimated by TIGAR (DPR), PrediXcan (Elastic-Net), and FUSION (best model) by ACAT.

### PPI network and pathway analysis

To investigate the PPI network among the identified PWAS risk genes of AD dementia, we used STRING^42, 43^, a website for PPI network analysis which integrates public data sources of protein interaction information and investigates the connectivity network of proteins. Proteins are considered as nodes in the PPI network. Protein-protein edges represent the predicted functional associations, colored differently to indicate seven categories –– co-expression, text-mining, experiments (biochemical/genetic data), databases (previously curated pathway and protein complex information), gene co-occurrence, gene fusion and neighborhood. Gene co-occurrence, fusion, and neighborhood represent association predictions are based on whole-genome comparisons.

STRING also enables gene-set enrichment analysis with respect to Gene Ontology (GO) annotations^62^, KEGG pathways^63^, Reactome pathways^64^, and UniProt keywords^65^. Enrichment analysis is to detect GO terms, pathways, or UniProt keywords that are significantly enriched with genes in the network versus random genes, adjusting for FDR. STRING also shows significant association between input proteins and human phenotypes based on Monarch Initiative^66^, a system to organize genotype-phenotype connections from multiple data sources.

### Test causal and pleiotropy effects

We used the PMR-Egger^18^ tool to test the vertical pleiotropy effect (i.e., causal mediation effect) between the protein abundance level of PWAS risk genes and AD dementia, while controlling for horizontal effect. The Multi-SNP-based SMR^59, 67^ software and its accompanying heterogeneity in dependent instruments (HEIDI) were also implemented to test the causal effect, using multiple *cis*-pQTL SNPs. Reference LD derived from ROS/MAP WGS data were used to implement both PMR-Egger and SMR-HEIDI methods. Test cis-SNPs were pruned according to their LD correlation matrix. The threshold of R^2^ = 0.95 was used for implementing PMR-Egger and R^2^ = 0.99 was used for implementing SMR-HEIDI. For each protein-coding gene, only SNPs that have nominal eQTL p-value < 0.05 were tested by SMR-HEIDI.

### TWAS-O of AD dementia

TWAS p-values were first obtained by using reference transcriptomic data of DLPFC tissue from ROS/MAP samples and GWAS summary data of AD dementia by TIGAR and PrediXcan, and then used to derive TWAS-O p-values by ACAT. The same GWAS summary data of AD dementia were used in both PWAS-O and TWAS-O analysis in this study. The reference transcriptomic data include 465 ROS/MAP samples with both gene expression data for DLPFC tissue (by RNA sequencing) and WGS genotype data profiled. Gene expression data in the format of Transcripts Per Million (TPM) were log2 transformed and adjusted for age at death, sex, postmortem interval, study (ROS or MAP), batch effects, RNA integrity number scores, estimated cell type proportions (with respect to oligodendrocytes, astrocytes, microglia, neurons), top five genotype PCs, and top probabilistic estimation of expression residuals (PEER) factors by linear regression models. The cell type proportions were estimated by using CIBERSORT pipeline^68^ with single-cell RNA-seq transcriptome profiles from human brain tissues as the reference^69^ to de-convolute bulk RNA-seq data^70^. Only genes with >0.1 TPM in ≥10 samples were considered. Cis-SNPs within a 1MB window of each gene that met the MAF threshold >1% and Hardy-Weinberg Equilibrium (HWE) threshold of p-value>10^-5^ were used in the imputation model as predictors. Only genes with an average 5-fold cross validation R^2^ ≥ 0.005 in the gene expression imputation models were analyzed in TWAS.

## Data Availability

All data produced in the present work are contained in the manuscript

https://doi.org/10.7303/syn3219045

## Acknowledgements

We are thankful to all participants and the staff at the Rush Alzheimer’s Disease Center for making the proteomics and WGS data of ROS/MAP samples available to the research community. This work was supported by the National Institute of Health R35GM138313 (T.H., R.P., Q.D., J.Y.). The ROS/MAP studies are supported by P30AG10161, K01AG054700, R01AG15819, R01AG17917, R01AG56352; the Illinois Department of Public Health; and the Robert C. Borwell Endowment Fund. The funding organizations had no role in the design or conduct of the study; collection, management, analysis, or interpretation of the data; or preparation, review, or approval of the manuscript.

## Author contributions

T.H. conducted data analysis and drafted the manuscript. J.Y. conceptualized and led the project, and edited the manuscript. R.P. and Q.D. contributed to data analysis. A.S.B., S.T., D.A.B., N.T.S. contributed to ROS/MAP data and edited the manuscript. M.P.E consulted data analysis results and edited the manuscript.

## Data availability

Proteomic, transcriptomic, and WGS data of ROS/MAP samples used in this manuscript are available from Synapse (https://doi.org/10.7303/syn3219045). These data are available via the AD Knowledge Portal (https://adknowledgeportal.org), subject to requirements for data access and data attribution as provided in https://adknowledgeportal.org/DataAccess/Instructions. GWAS summary data of AD dementia is available from https://ctg.cncr.nl/software/summary_statistics. Results of the pQTL weights and PWAS summary data generated in this study will be deposited to Synapse after publication of this work. Instructions for implementing PWAS-O are available through PWAS-O Github page (https://github.com/tingyhu45/PWAS-O).

## Ethics declarations

Both MAP and ROS studies were approved by the Institutional Review Board of Rush University Medical Center. All individual-level data analyzed in this work were de-identified. Written informed consent was obtained from all study participants as was an Anatomical Gift Act for organ donation. ROSMAP resources can be requested at www.radc.rush.edu.

## Supplementary information

The supplementary file contains 2 supplementary tables and 8 supplementary figures.

