## Supplementary Materials for "Omnibus proteome-wide association study (PWAS-O) identified 43 risk genes for Alzheimer’s disease dementia"

### Supplementary information

**Table S1. Significant PWAS-O risk genes with overlapped test SNPs with the nearby top significant PWAS-O risk gene as in Table 2.**
Missing P-values denoted by dashes are due to either CV $R^2$< 0.005 for the corresponding protein abundance imputation models, or no trained protein abundance imputation model by PrediXcan.

| Gene | CHR | PWAS-O P-value | TIGAR P-value | PrediXcan P-value | FUSION P-value |
|---|---|---|---|---|---|
| C4A$^{b,c}$ | 6 | $4.02 \times 10^{-5}$ | - | - | $4.02 \times 10^{-5}$ |
| MBLAC1$^{b,c}$ | 7 | $1.81 \times 10^{-4}$ | $7.31 \times 10^{-5}$ | $3.49 \times 10^{-4}$ | $6.67 \times 10^{-1}$ |
| ACTL6B$^{b}$ | 7 | $6.52 \times 10^{-5}$ | $6.52 \times 10^{-5}$ | - | - |
| FNBP4$^{b,c}$ | 11 | $3.06 \times 10^{-5}$ | $3.06 \times 10^{-5}$ | - | - |
| FAM111B$^{b}$ | 11 | $1.60 \times 10^{-4}$ | $1.60 \times 10^{-4}$ | - | - |
| PRPF19$^{b}$ | 11 | $2.45 \times 10^{-5}$ | $1.22 \times 10^{-5}$ | - | $1.12 \times 10^{-1}$ |
| PACSIN3 | 11 | $8.19 \times 10^{-5}$ | $8.19 \times 10^{-5}$ | - | - |
| TMEM132A$^{b,c}$ | 11 | $8.50 \times 10^{-6}$ | $8.50 \times 10^{-6}$ | - | - |
| KIF18B$^{b,c}$ | 17 | $1.25 \times 10^{-4}$ | $2.94 \times 10^{-4}$ | - | $7.92 \times 10^{-5}$ |
| HEXIM1$^{b}$ | 17 | $1.82 \times 10^{-4}$ | $1.88 \times 10^{-1}$ | - | $9.09 \times 10^{-5}$ |
| PVR$^{a,b,c}$ | 19 | $8.48 \times 10^{-9}$ | $8.48 \times 10^{-9}$ | - | - |
| PVRL2$^{a,b,c}$ | 19 | $2.81 \times 10^{-4}$ | $2.81 \times 10^{-4}$ | - | - |
| TOMM40$^{b,c}$ | 19 | $4.43 \times 10^{-8}$ | $4.43 \times 10^{-8}$ | - | - |
| APOC1$^{a,b,c}$ | 19 | $1.20 \times 10^{-4}$ | $1.20 \times 10^{-4}$ | - | - |
| APOC2 $^{a,b,c}$ | 19 | $2.35 \times 10^{-22}$ | $2.35 \times 10^{-22}$ | - | - |
| CKM$^{a,c}$ | 19 | $1.67 \times 10^{-7}$ | $1.67 \times 10^{-7}$ | - | - |
| ERCC2$^{b,c}$ | 19 | $1.01 \times 10^{-39}$ | $3.37 \times 10^{-40}$ | $5.29 \times 10^{-35}$ | $3.01 \times 10^{-20}$ |
| ERCC1$^{a,b,c}$ | 19 | $5.40 \times 10^{-15}$ | $5.40 \times 10^{-15}$ | - | - |
| RTN2$^{b,c}$ | 19 | $1.33 \times 10^{-24}$ | $1.33 \times 10^{-24}$ | - | - |
| PPM1N$^{b,c}$ | 19 | $3.14 \times 10^{-6}$ | $2.01 \times 10^{-4}$ | $6.90 \times 10^{-1}$ | $1.05 \times 10^{-6}$ |
| OPA3$^{a,b,c}$ | 19 | $1.85 \times 10^{-24}$ | $1.85 \times 10^{-24}$ | - | - |
| EML2$^{b,c}$ | 19 | $4.88 \times 10^{-13}$ | $4.88 \times 10^{-13}$ | - | - |
| DMWD$^{b,c}$ | 19 | $2.37 \times 10^{-20}$ | $1.96 \times 10^{-1}$ | $2.19 \times 10^{-20}$ | - |
| SYMPK$^{a,b,c}$ | 19 | $1.01 \times 10^{-5}$ | $5.04 \times 10^{-6}$ | $9.50 \times 10^{-1}$ | - |

*a* Known GWAS risk genes of AD.
*b* Known TWAS risk genes of AD dementia, or test region overlapped with TWAS risk genes.
*c* Significant mediated causal genetic effects by PMR-Egger or SMR.

**Table S2. PWAS-O risk genes with test region overlapped with TWAS-O risk genes of AD dementia.**

34 out of 43 PWAS risk genes were also identified by TWAS-O with ROS/MAP transcriptomic reference panel of DLFPC tissue and the same GWAS summary data of AD dementia.

| Gene | CHR | PWAS-O P-value | TWAS-O P-value |
|---|---|---|---|
| ERCC3 | 2 | $2.91 \times 10^{-4}$ | $8.50 \times 10^{-7}$ |
| HSPA1B | 6 | $2.64 \times 10^{-9}$ | $1.07 \times 10^{-5}$ |
| TULP1 | 6 | $2.88 \times 10^{-4}$ | $7.94 \times 10^{-5}$ |
| C4A | 6 | $4.02 \times 10^{-5}$ | $5.90 \times 10^{-5}$ |
| TRIM4 | 7 | $3.72 \times 10^{-5}$ | $5.30 \times 10^{-5}$ |
| MBLAC1 | 7 | $1.81 \times 10^{-4}$ | $1.04 \times 10^{-4}$ |
| AGFG2 | 7 | $9.86 \times 10^{-8}$ | $8.03 \times 10^{-5}$ |
| ACTL6B | 7 | $6.52 \times 10^{-5}$ | $8.03 \times 10^{-5}$ |
| ZYX | 7 | $1.39 \times 10^{-5}$ | $4.12 \times 10^{-5}$ |
| FAM111B | 11 | $1.60 \times 10^{-4}$ | $6.05 \times 10^{-7}$ |
| MRPL16 | 11 | $8.66 \times 10^{-15}$ | $1.32 \times 10^{-4}$ |
| CCDC86 | 11 | $2.83 \times 10^{-9}$ | $4.89 \times 10^{-10}$ |
| PRPF19 | 11 | $2.45 \times 10^{-5}$ | $4.89 \times 10^{-10}$ |
| TMEM132A | 11 | $8.50 \times 10^{-6}$ | $4.89 \times 10^{-10}$ |
| ATPAF2 | 17 | $8.20 \times 10^{-5}$ | $1.23 \times 10^{-4}$ |
| KIF18B | 17 | $1.25 \times 10^{-4}$ | $5.57 \times 10^{-5}$ |
| DCAKD | 17 | $6.19 \times 10^{-5}$ | $5.57 \times 10^{-5}$ |
| HEXIM1 | 17 | $1.82 \times 10^{-4}$ | $5.57 \times 10^{-5}$ |
| PVR | 19 | $8.48 \times 10^{-9}$ | $7.45 \times 10^{-10}$ |
| PVRL2 | 19 | $2.81 \times 10^{-9}$ | $3.55 \times 10^{-15}$ |
| TOMM40 | 19 | $4.43 \times 10^{-8}$ | $3.55 \times 10^{-15}$ |
| APOC1 | 19 | $1.20 \times 10^{-4}$ | $3.55 \times 10^{-15}$ |
| APOC2 | 19 | $2.35 \times 10^{-22}$ | $3.55 \times 10^{-15}$ |
| CLPTM1 | 19 | $1.44 \times 10^{-70}$ | $3.55 \times 10^{-15}$ |
| CKM | 19 | $1.67 \times 10^{-7}$ | $7.68 \times 10^{-7}$ |
| ERCC2 | 19 | $1.01 \times 10^{-39}$ | $7.68 \times 10^{-7}$ |
| ERCC1 | 19 | $5.40 \times 10^{-15}$ | $7.68 \times 10^{-7}$ |
| RTN2 | 19 | $1.33 \times 10^{-24}$ | $7.68 \times 10^{-7}$ |
| PPM1N | 19 | $3.14 \times 10^{-6}$ | $7.68 \times 10^{-7}$ |
| OPA3 | 19 | $1.85 \times 10^{-24}$ | $7.68 \times 10^{-7}$ |
| EML2 | 19 | $4.88 \times 10^{-13}$ | $6.36 \times 10^{-5}$ |
| DMWD | 19 | $2.37 \times 10^{-20}$ | $6.36 \times 10^{-5}$ |
| SYMPK | 19 | $1.01 \times 10^{-5}$ | $6.36 \times 10^{-5}$ |
| SPHK2 | 19 | $1.76 \times 10^{-4}$ | $2.09 \times 10^{-5}$ |

**Fig. S1. Quantile-Quantile (QQ) plots for TIAGR (DPR), PrediXcan, FUSION and PWAS-O under null simulations.**

Phenotype data were simulated from $N(0,1)$. Protein expression were generated with $h_p^2 = 0.05$ and $p_{causal} = 0.001$. pQTL weights were permuted $2 \times 10^3$ times for 500 genes to perform a total of $10^6$ null simulations of PWAS.

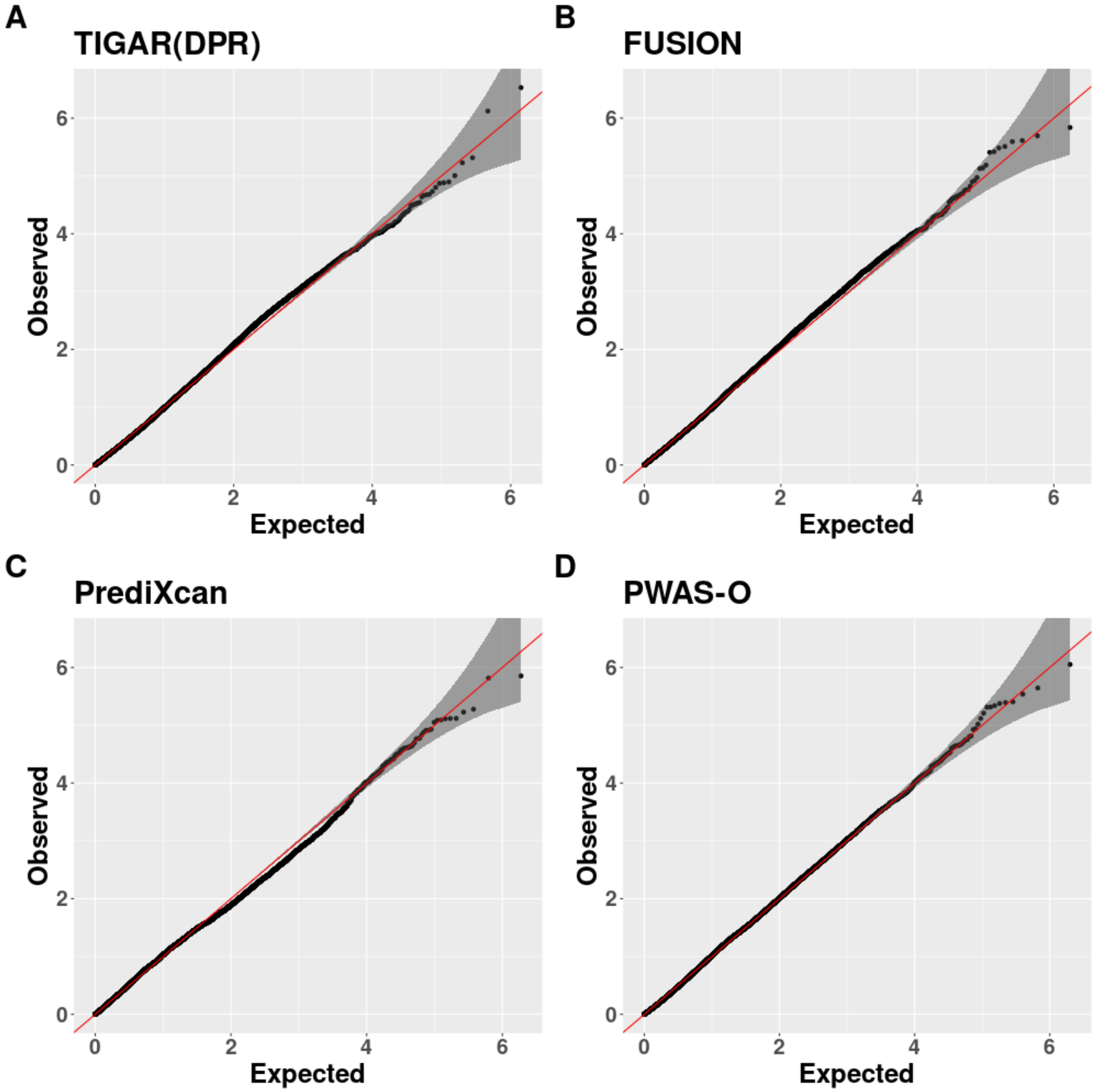

**Fig. S2. Quality control by principal component analysis for human brain protein abundances.** Top 2 principal components (PCs) of the protein abundance data of 400 samples were plotted in panel A, with the red line representing four times of the standard deviations from the mean of PC1 and blue representing four times of the standard deviations from the mean of PC2. After excluding outlier samples that fell outside of the red and blue lines in panel A, the top 2 PCs with remain samples were plotted in panel B. Similarly, after excluding outlier samples that fell outside of the red and blue lines in panel B, the top 2 PCs with remain samples were plotted in panel C. Five samples were identified as outliers and excluded from PWAS-O analyses.

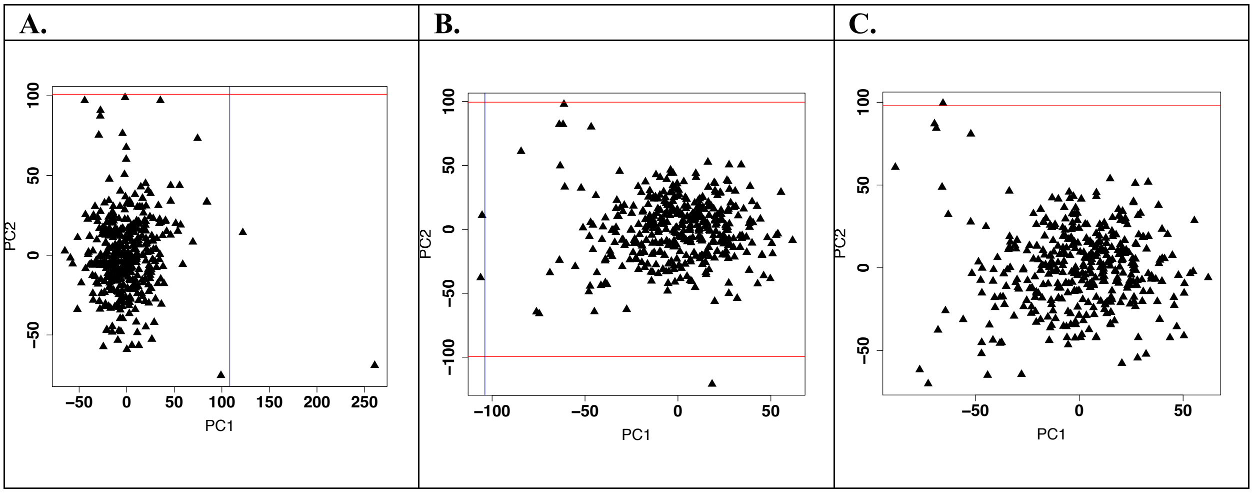

**Fig. S3. Histogram plots of 5-fold CV $R^2$ for trained protein abundance imputation models by TIGAR, PrediXcan, and FUSION, using ROS/MAP proteomic reference data.**

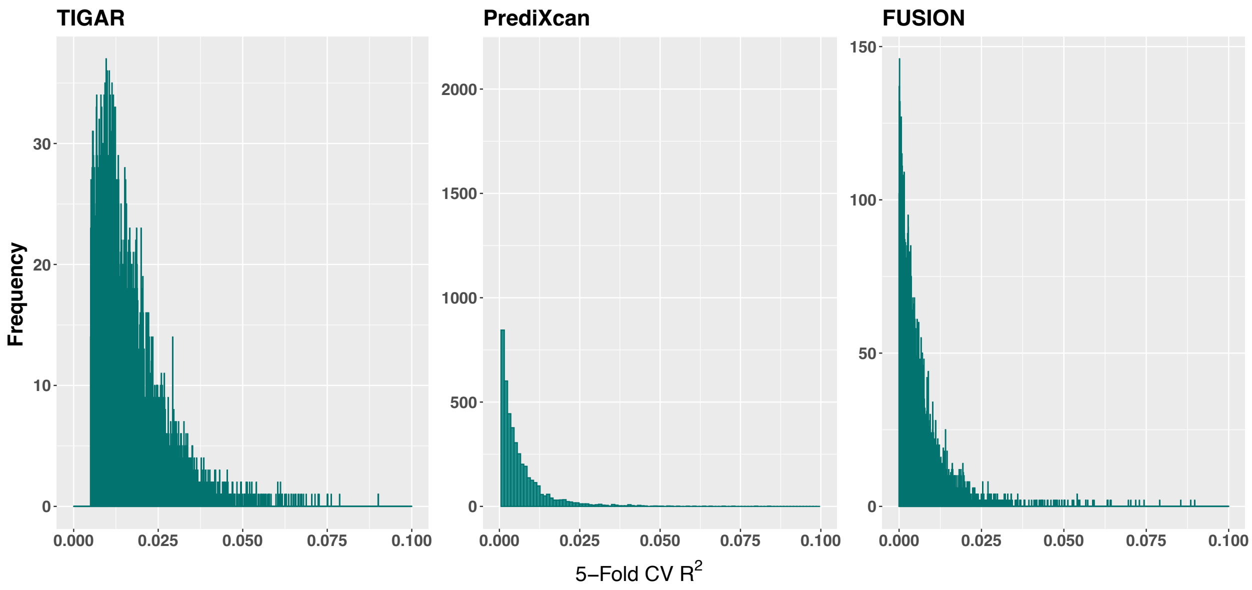

**Fig. S4. Manhattan plots of PWAS results (FDR q-values) for studying AD dementia by TIGAR (A), PrediXcan(B), and FUSION(C).**
The -log10(q-values) were plotted in y-axis, and -log10(0.05) was plotted as the dashed horizontal line. Independently significant genes are labeled in the plots.

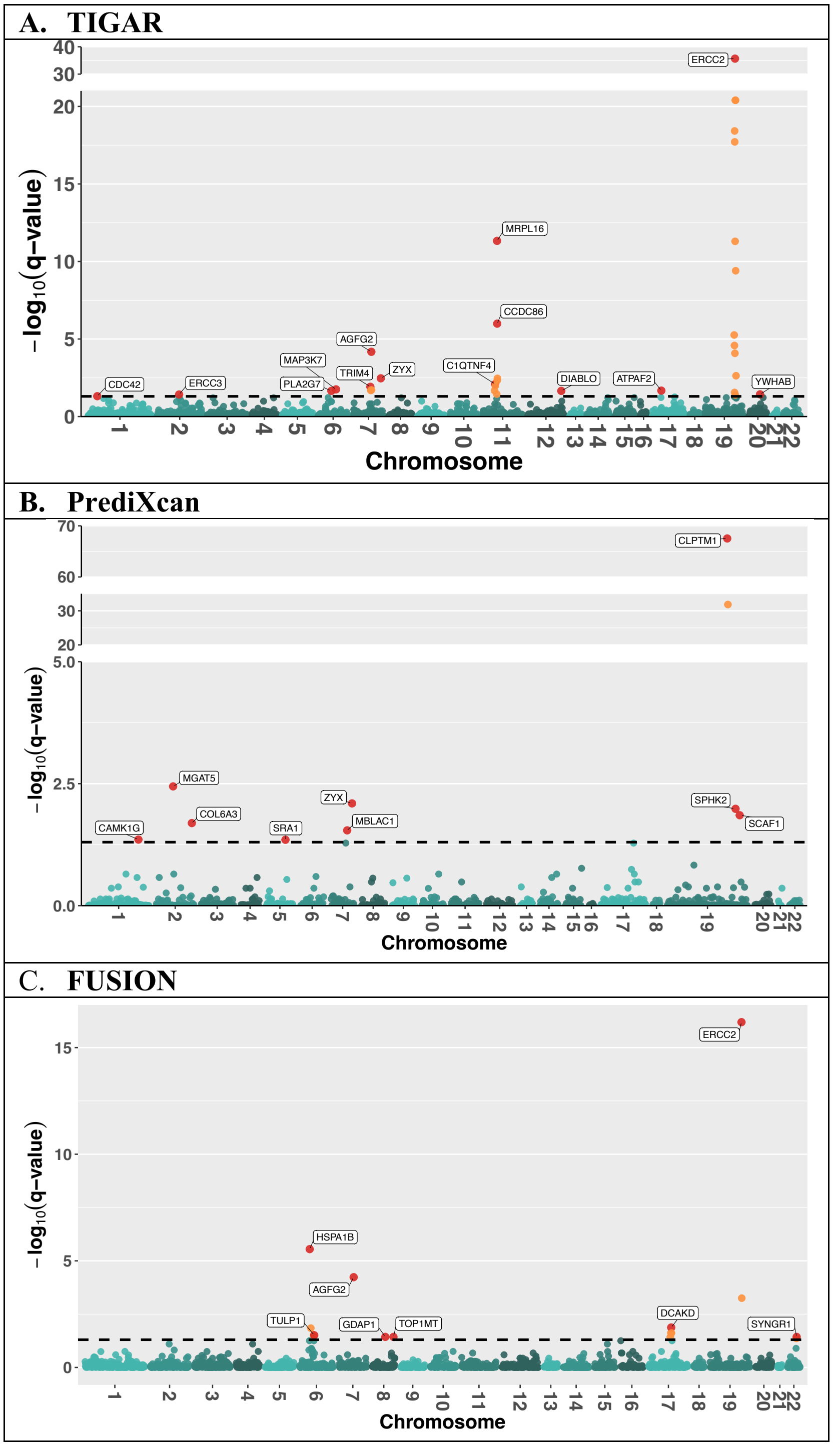

**Fig. S5. Quantile-quantile plots for PWAS-O (A), and individual PWAS by TIGAR (B), PrediXcan (C) and FUSION(D) of AD dementia.**
Genomic control factors were obtained as $\lambda = 1.14$ for PWAS-O of AD, $\lambda = 1.11$ for TIGAR/DPR, $\lambda = 1.08$ for PrediXcan, and $\lambda = 1.25$ for FUSION.

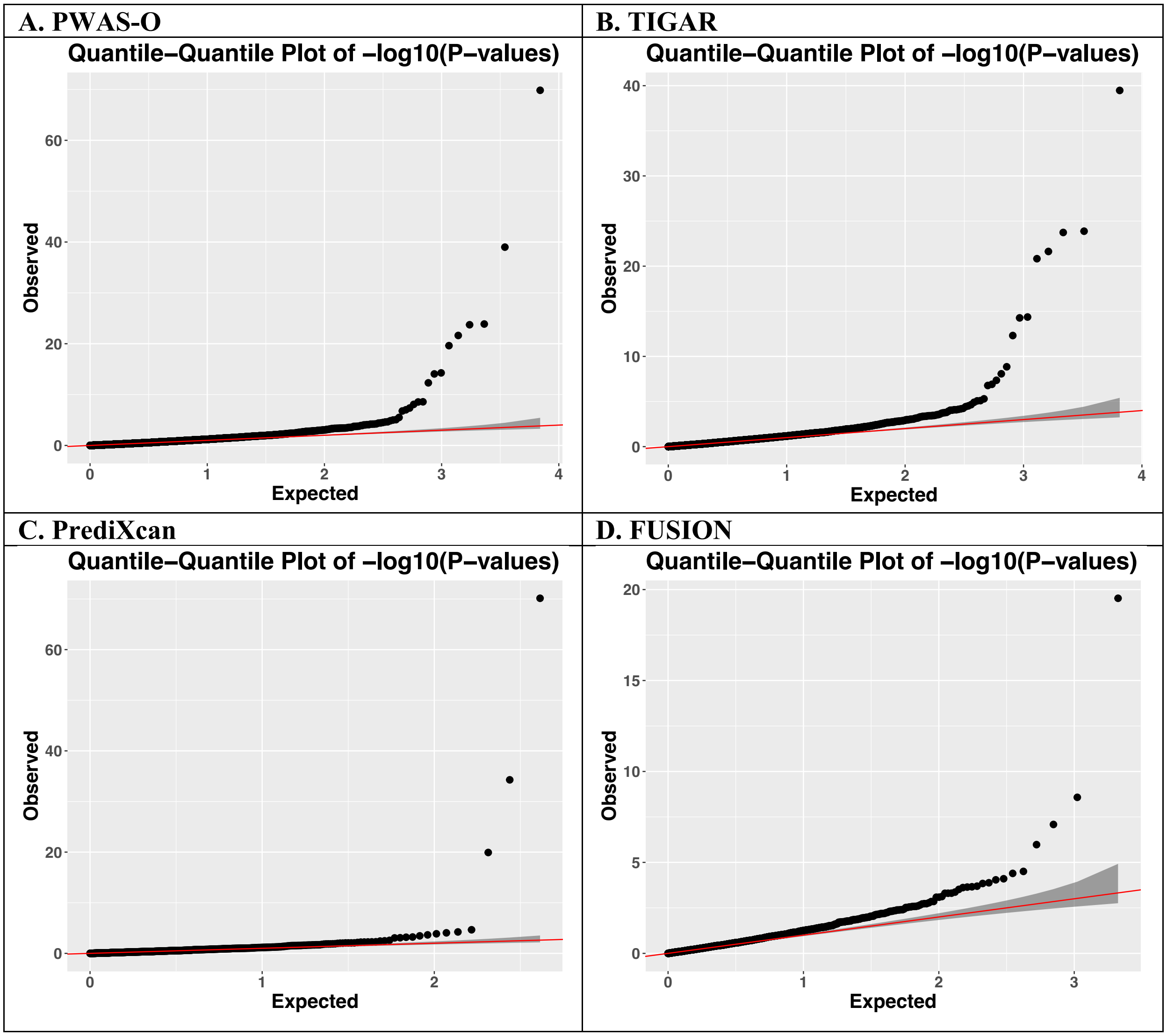

**Fig. S6. Venn diagram of the number of shared risk genes identified by PWAS-O and the individual PWAS methods for AD dementia.**

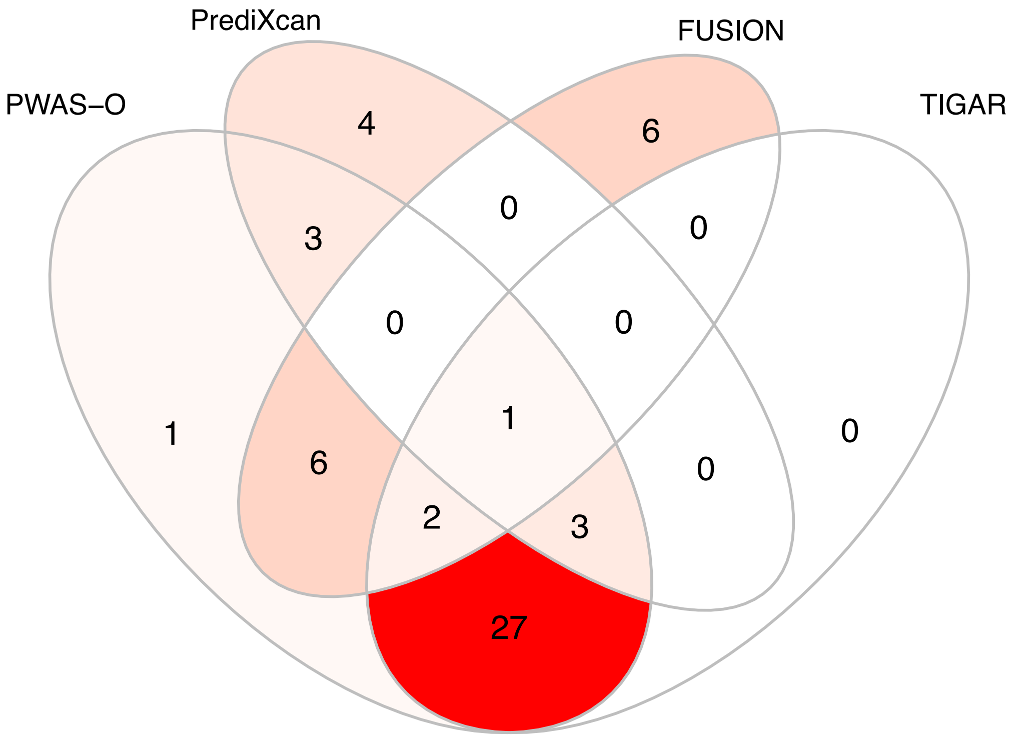

**Fig. S7. Estimated pQTL weights by TIGAR (DPR) and PrediXcan (Elastic-Net) for *MRPL16* (A) and *MBLAC1* (B*).***

The pQTL weights were plotted in the y-axis for all test SNPs in the test gene, with color coded with -log10 (GWAS p-value). Test SNPs with GWAS p-value $<10^{-5}$ were plotted in yellow color. The significance of gene *MRPL16* was driven by TIGAR as more test SNPs with non-zero pQTL weights by TIGAR were colocalized with GWAS significant SNPs (A). Gene *MBLAC1* was found significant by PWAS using both TIGAR and PrediXcan, for having non-zero pQTL weights for SNPs colocalized with significant GWAS signals (B).

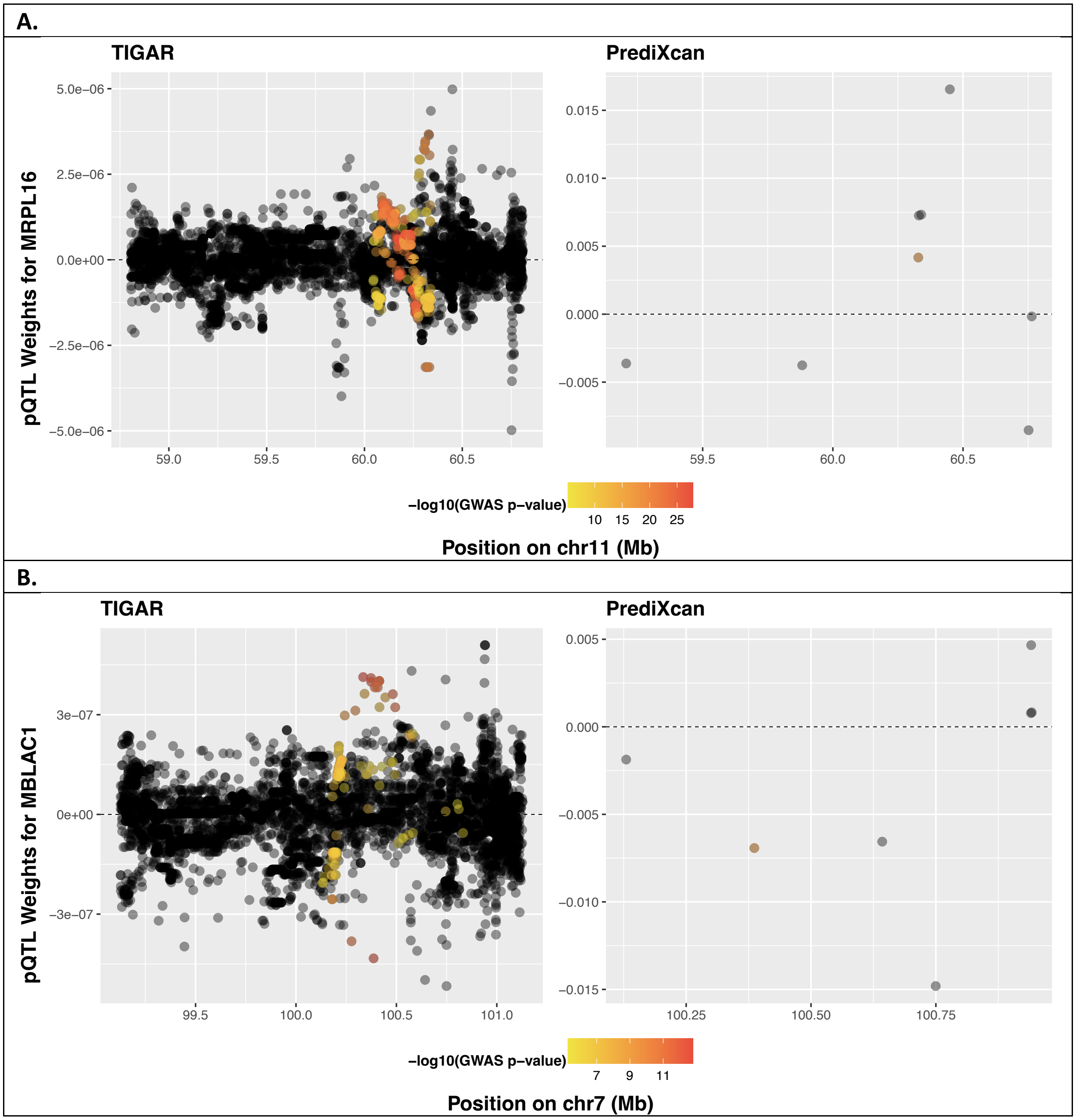

**Fig. S8. Venn diagram of the number of shared risk genes by TWAS-O and PWAS-O.**
A total of 34 genes were either commonly identified by TWAS-O and PWAS-O or had overlapped test region (within 1MB of the transcription starting sites and termination sties of the test protein coding gene).

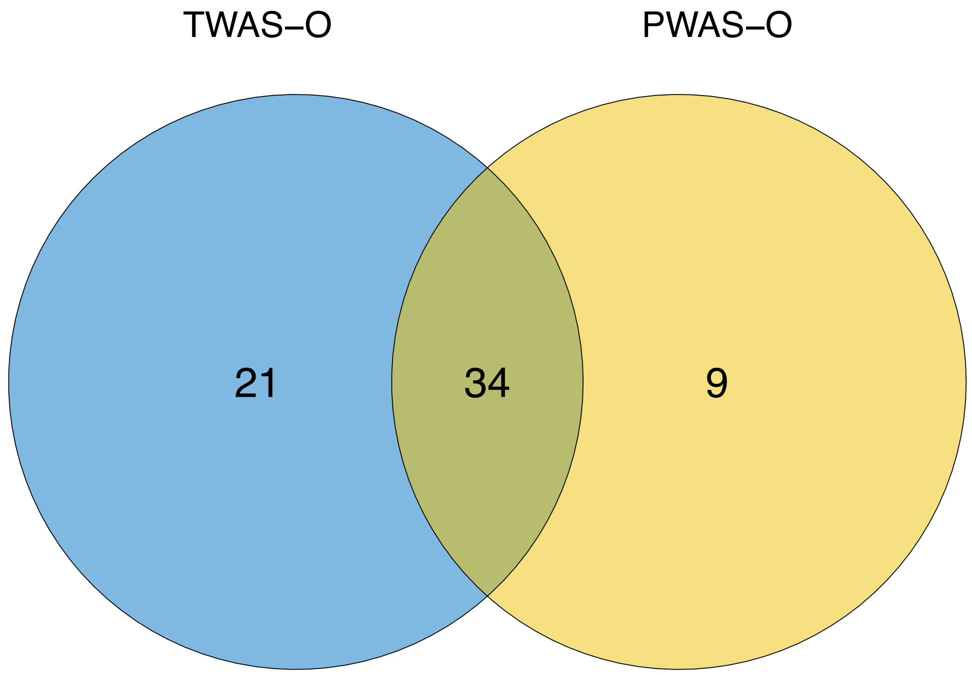
